# Acceptability, willingness to use and preferred distribution models of oral-based HIV self-testing kits among key and priority populations enrolled in HIV pre-exposure prophylaxis clinics in central Uganda. A mixed-methods cross-sectional study

**DOI:** 10.1101/2022.07.27.22278119

**Authors:** John Bosco Junior Matovu, Roy William Mayega, Sylivia Nalubega, Jayne Byakika-Tusiime

## Abstract

Key Populations (KPs) and Priority Populations (PPs) taking Pre-Exposure Prophylaxis (PrEP) for HIV prevention require routine HIV testing every after three months. HIV self-testing could be an alternative testing approach for these population categories. We assessed the acceptability of oral-based HIV Self-Testing (HIVST) among Key and priority Populations taking PrEP in central Uganda. A mixed methods cross-sectional study was conducted on 367 key and priority populations attending Pre-Exposure Prophylaxis clinics in central Uganda, from May to August 2018. KPs and PPs were introduced to the Oraquick HIV self-testing kit as an option for HIV testing during their routine visit to the PrEP clinic. A demonstration of how to perform HIV self-testing was conducted using an Oraquick demonstration video and leaflet inserts after which, respondents were asked to choose between HIVST and the conventional facility blood-based HIV testing. Those willing to use the oral kit were asked to voluntarily consent, were provided with an Oraquick HIVST kit and were assisted to perform the test. Quantitative data were presented as proportions for each outcome variable. Thematic analysis was performed to explore factors that promote and inhibit HIVST. HIV self-testing acceptability; defined as the proportion of those who performed an HIVST test among those approached was 99.5% (365/367). Using an oral fluid-based kit “Oraquick” was reported to be free of pain, convenient, easy to use and time saving hence preferred over other HIV testing modalities. A multimodal approach to distributing HIV self-testing kits was suggested by respondents. Oral-based HIV self-testing is highly acceptable among key and priority populations taking Pre-Exposure Prophylaxis and can be adopted as an alternative to the conventional routine three monthly facility-based provider dependent HIV screening. Kits’ distribution may employ several models. Majority of key populations would afford the kits at a cost of not more than 1.4USD if not provided free of charge.

## Introduction

Globally, an estimated 84% of all people living with Human Immunodeficiency Virus (HIV) know their status [1]. As the world prepares to end AIDS by 2030, the remaining undiagnosed people living with HIV need to be identified, initiated and maintained on treatment to achieve viral suppression [2]. In Uganda, most of the new HIV infections occur in Key Populations (KPs) and Priority Populations(PPs) who include sex workers, fisher folks, long distance truck drivers, uniformed service personnel, men who have sex with men (MSM), and boda-boda taxi-men [3]. Pre-Exposure Prophylaxis (PrEP) was introduced as a biomedical intervention to reduce the risk of HIV acquisition among people with ongoing risk of HIV acquisition such as KPs and PPs. Commonly used PrEP medication is a formulation of a fixed dose combination drug consisting of Tenofovir (TDF) and Emtricitabine (FTC), taken daily as long as the risk of HV acquisition remains. However, injectable PrEP formulations such as long acting Cabotegravir have been developed and provide better adherence on PrEP hence better prevention outcomes. People taking PrEP are required to test for HIV every three months to ascertain their HIV negative status so as to continue with prevention services. Currently, there is only one existing testing method for routine follow up testing of KPs on PrEP: that is, blood-based test either at the clinic (facility) or at community outreach centers.

HIV Self Testing (HIVST) is an HIV testing approach where a person collects his or her own specimen (oral fluid or blood), performs an HIV test and interprets the result, often in a private setting, either alone or with someone he or she trusts [4]. Providing a variety of HIV testing options is a good strategy for increasing HIV testing uptake and would potentially improve HIV testing among KPs and PPs leading to early linkage to post-test services [5].

Studies conducted on HIV self-testing worldwide have shown high but varying levels of acceptability across age, gender and sub-populations [6-11].

Among PrEP users, a study conducted in Kenya on the feasibility and acceptability of HIV self-testing revealed that 90% (N=2400) of HIV negative respondents in sero-discordant relationships on PrEP accepted to use self-testing kits [12] but discordant couples account for only about 2% of the entire clients on PrEP in Uganda.

Barriers to acceptability of HIVST have been reported and include unease of use, cost of the test kits, and lack of accessibility to professional support after self-testing [10, 13] and as noted by Figueroa et al. [11], respondents expressed concerns over lack of counselling before self-testing and accuracy of results since the test is not performed by a health professional. For effective implementation and optimisation of the outcomes of HIVST, these barriers and concerns need to be adequately addressed.

There is limited literature about studies conducted to determined acceptability of HIV self-testing using an oral based kit among KPs and PPs who are taking PrEP. We aimed to determine the proportion of Key and Priority Populations taking Pre-Exposure Prophylaxis for HIV in central Uganda who accept oral-based HIV Self-Testing, the preferred HIV testing method for routine testing while on PrEP, their willingness to pay for the kits and the amount they are willing to pay and to establish the preferred HIV Self-Testing kit distribution approaches/models. Our study builds on existing literature about HIV self-testing and contributes a new dimension of using HVST as a favourable HIV testing approach among KPs and PPs on PrEP during routine three monthly follow up testing.

## Methods

### 2.1 Study design and setting

This cross-sectional study employed both quantitative and qualitative research methods and was conducted between May and August 2018 at two clinics in Uganda where PrEP was being offered: Most At Risk Population Initiative (MARPI) clinic in Kampala city and Kasensero HC II in Rakai district.

The study targeted all people categorized as key populations [Men who have sex with Men (MSM), People who inject drugs (PIDs), Female sex workers (FSWs)] and priority populations [(Fisher folks, Adolescent girls and young women (AGYWs) and negative people in discordant sexual relationships)] who had been enrolled and active on PrEP at both PrEP clinics by December 2017. Two emancipated minors aged 17 years belonging to the adolescent girls and young women category were considered for the study and written informed consent was obtained from each of their guardians before enrolment[14, 15]. We excluded respondents who had ever performed HIV self-testing prior to the study and those who were unable to provide consent on their own either due to ill health, altered mental state or any other reason.

We estimated the sample size using the Kish Leslie formula for cross-sectional studies for a single sample proportion for a categorical outcome.

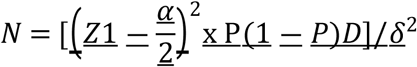

N= Calculated sample size

P= assumed sample proportion that accepts HIVST estimated at 85% (P=0.85).

The estimated acceptability of 85% was based on assumptions that HIVST was new in the study population and there was no in-country data on acceptability of HIVST in the study population, hence we anticipated the below that of Nangendo et al. [6] and Ngure et al. [12], hence we expected a lower acceptance rate of HIVST among study participants compared to the general population

1-P = The probability of not accepting the HIVST

Z1-α/2 = Standard normal deviate at 95% confidence interval (z=1.96)

δ =acceptable margin of error acceptability of 5%.

D=Design effect of 1.5 was included in the sample size estimate because our respondents were sampled from two different clinics (Clusters) purposively selected, and further still, respondents were being reviewed at each of those clinics serving as clusters. An estimated dropout rate of 20% was considered because HIVST was a new intervention among respondents.

Due to logistical reasons and the dynamic nature of the study population, we employed a consecutive convenience sampling strategy. Potential participants were screened for eligibility as they came for their clinic appointments until the desired sample size was achieved for each clinic to make a total of 367 respondents.

The primary outcome (dependent) variable for the study was acceptability to use HIV self-testing at this particular follow up visit. Acceptability was defined as the proportion of respondents who used the oral HIVST kit out of those reached with the HIVST. Secondary outcomes were: preferred method of HIV testing during routine follow up monitoring, willingness to pay for the HIVST kit, amount to pay for an HIVST kit and the preferred model of distributing HIVST kits. Social, demographic and economic data including age, gender, KP/PP category, education level, occupation, marital status and religion, and level of income were assessed

### 2.2 Data collection and management

Data collection took place between May and August 2018. Quantitative data was collected using an interviewer administered structured questionnaire in English language and questions were directly translated in the preferred local language during the interview session for respondents who did not understand English. In-depth individual interviews were used to collect qualitative data after collection of the quantitative data.

Respondents were informed of the availability of HIVST and the standard HIV testing services at the clinic, followed by a demonstration of HIV self-testing procedure by research assistants using a demonstration video and leaflet inserts in the HIVST pack. The demonstration involved three steps: preparing for oral fluid sample collection, using the kit to test for HIV, and interpreting test results. Respondents were asked if they were willing to perform a self-test and those willing were asked to voluntarily consent after which, they were provided with an Oraquick HIVST kit to perform self-testing (accepting) and interpret the results. As a quality assurance measure, research assistants supervised the self-testing procedure. Respondents who tested HIV positive (five in number) on self-testing were subjected to a confirmatory test using the national HIV testing algorithm. Respondents who tested HIV positive on confirmatory testing (four in number) did not continue with the questionnaire.

### 2.3 Data analysis

Data was analyzed using STATA 14.2 version. Univariate analysis was used to calculate the descriptive statistics for each of the variables. The categorical variables were summarized as proportions, while the continuous variables as means and standard deviation.

We measured HIV testing kit type preference by computing the proportion of respondents who would prefer to use one of the three HIV testing choices available, that is; a self-testing kit, a blood based self-testing kit if introduced in future or using the conventional health facility based-health worker provided HIV test. We determined HIVST kit distribution model preference by computing the proportion of respondents who would prefer any of the available testing kit distribution models. We determined willingness to pay by computing the proportion of respondents who reported they would pay for an HIVST if it was at a cost. The amount one was willing pay was captured in Uganda shillings (UGX) and converted in USD equivalent during analysis.

The qualitative study employed an in-depth interview technique to gather data from participants and followed the quantitative data collection. We interviewed 20 participants with effort to ensure variation between KP categories, gender and testing site. Purposive sampling was used to select the participants and the principle of data saturation (where no new insights emerge from the data) was used to guide participant enrolment into the study. Recorded information was transcribed verbatim and important statements were identified and extracted. The thematic analysis approach was employed for data analysis and results were presented in a narrative form alongside and to support the quantitative results.

### 2.4 Power calculations

Based on post Clopper-Pearson confidence interval formula, with a sample size of 367, a width of 0.5 and a sample proportion, we were powered to detect the observed primary outcome with *>*90% power.

### 2.5 Ethics Approval and Consent to participate

The study protocol was reviewed and approved by the Busitema University Faculty of Health Sciences Higher Degrees and Research Committee and the Mbale Regional Referral Hospital Research and Ethics Committee (Ref No. MRRH-REC-IN-COM 024/2018). All respondents provided written informed consent and were free to opt out of the study at any stage of the study.

## Results

### 3.1 Participants

As indicated in figure 1, a total of three hundred sixty-seven (367) respondents were reached, one respondent opted out and preferred not to be interviewed for personal reasons. Out of the 366 respondents who were willing to perform HIV self-testing, 365 performed the test. Of the 365 respondents who performed a self-test, five respondents tested HIV positive. Both the negative (360) and the positive (five) respondents were subjected to national HIV testing algorithm and four were confirmed HIV positive. All positive participants were linked to Anti-Retroviral Therapy (ART) clinics for initiation on treatment. The fifth respondent who tested negative on the national HIV testing algorithm was considered HIV negative. For the qualitative component, twenty interviews were conducted, eleven males and nine females with an age range of 19-40. Twelve participants were from MARPI Mulago while eight were from Kasensero HC III clinics. Overall, for the qualitative study, there were five female sex workers, three people who use and inject drugs, three MSM, six HIV negative people in sero-discordant sexual relationships, two young people and one fisherman. Four main themes emerged from the qualitative study which were generally aligned to the study aim. These were: acceptability of HIVST, choice of an HIV testing approach, preferred kit distribution models and willingness to pay for the test.

**Figure 1:**
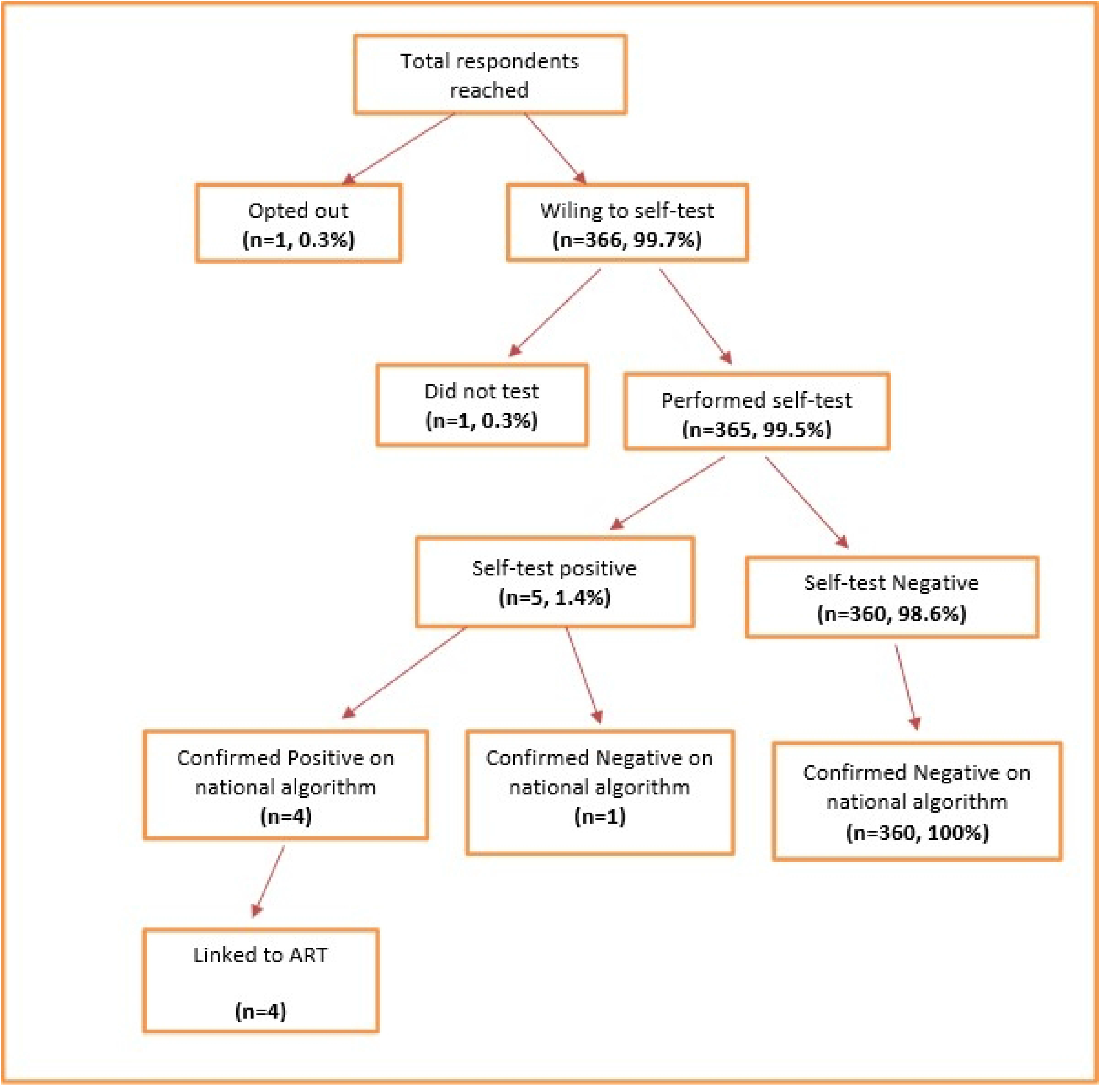
Flow diagram for client participation.

### 3.2 Socio demographic and economic characteristics of participants

Three hundred sixty-six individuals were included in the study. The mean age was 28 (SD=7.5), the youngest participant was 17 years while the oldest at 54 years, 54.1% (n=198) were from urban settings, 49.2% (n=180) were males, 34.2% (n=125) were female sex workers and 48.4% (n=177) had attained up to primary education. About 34% (n=124) were married or cohabiting and more than half 52.5%, (n=192) were Catholics. Casual labor was the major source of income contributing to 62.6%. (n=229) as detailed in table 1 below.

**Table 1:** Socio demographic and economic characteristics of the study participants.

|  | **n (N=366)** | **%** |
| --- | --- | --- |
| **Site** |  |  |
| MARPI Mulago (Urban) | 198 | 54.1 |
| Kasensero (Rural) | 168 | 45.9 |
| **Age category** |  |  |
| 17-19 | 34 | 9.3 |
| 20-24 | 96 | 26.2 |
| 25-29 | 103 | 28.1 |
| 30-34 | 56 | 15.3 |
| 35-39 | 42 | 11.5 |
| 40-44 | 21 | 5.7 |
| 45 and above | 14 | 3.8 |
| **Sex** |  |  |
| Male | 180 | 49.2 |
| Female | 186 | 50.8 |
| **Education level** |  |  |
| Never went to school | 25 | 6.8 |
| Primary | 177 | 48.4 |
| Secondary | 141 | 38.5 |
| Tertiary institutions (none-degree) | 13 | 3.6 |
| University | 10 | 2.7 |
| **Marital status** |  |  |
| Single/Never Married | 117 | 32 |
| Married/Cohabiting | 124 | 33.9 |
| Divorced/Separated/widowed | 125 | 34.1 |
| **Religion** |  |  |
| Catholic | 190 | 51.9 |
| Protestant | 70 | 19.1 |
| Muslim | 72 | 19.7 |
| Pentecostal | 27 | 7.4 |
| **Key/Priority population category** |  |  |
| AGYW | 31 | 8.5 |
| Discordant Negative | 36 | 9.8 |
| Fisher Folks | 79 | 21.6 |
| FSW | 125 | 34.2 |
| MSM | 47 | 12.8 |
| PWID | 45 | 12.3 |
| Transgender | 3 | 0.8 |
| **Source of income** |  |  |
| Formally employed (Gov't/NGO/Private) | 24 | 6.6 |
| Casual labor | 229 | 62.5 |
| Self Employed | 84 | 23.0 |
| In school, not employed | 7 | 1.9 |
| Farmer | 22 | 6.0 |

### 3. Acceptability of HIV self-testing

Acceptability was computed as a proportion of respondents who performed HIV self-test out of those who were reached with HIV self-testing. We reached 367 participants and 365 performed HIV self-testing translating into an acceptability of 99.5%. One participant declined to take the kit and one other participant did not perform the self-test.

Participants felt HIVST was an approach that would greatly reduce obstacles associated with current approaches to HIV testing, citing that as opposed to the conventional HTS approaches, HIVST was time saving, cost effective, private and accessible ***(‘you move with your lab’)***. These factors were seen as motivations to use a self-test.

*“The other problem is having to line up, I may come but when I have no time for lining up and I go back without being tested. I will sit until the line is completed, yet here I will be alone and still go when I know whom I am” (M05, MSM)*.

The time factor was particularly an important concern for special categories of people, such as sex workers, who work during night and sleep during day hence find day time very valuable. Additionally, some participants felt HIVST was especially important for special groups who feel stigmatised when they go to health facilities. Particularly, FSWs and MSMs reported this concern and felt society has not accepted them and discriminates them, hence found HIVST more convenient.

> *Like for us (MSM), there are those who do not want to be identified. When they just want to be in hiding, but for us we bump on them. This method will be helpful when they come to know their status and take care accordingly, because they may say they have no time, and you offer them the test (M05, MSM)*.

Specifically, those who are likely to engage in unplanned sex e.g. those who use drugs and sex workers felt HIVST was very convenient, as they may need to test a sexual partner before engaging into sex, something not easy with facility based approaches. On the other hand, those who use and inject drugs stated that drugs can abruptly increase their sexual desire which can lead them to engage with irregular sexual partners. In this case, they will be safer by testing them first, hence the need for HIVST.

*“It is good because it is handy, any time when you have someone, you can use it with a customer (sex customer), because many will fear to go to the clinic. So it is safe for you to move with your own test” (M10, FSW)*.

> *I was excited as a person and I thought if I can have somewhere to find them I would not be worried, because like some of us who use drugs, you may be there high on drugs and you just pick up a woman, and by the time you come to your senses, you regret your actions, […]. But when you have that test, it becomes easy for you to test yourself and take caution to protect your-self (M01, PUWD)*.

In addition to testing one individual, HIVST was seen as being capable of engaging many more people (through peer recommendation), and thus increase the number of people who will become aware of their HIV status.

Many participants expressed that they trusted HIVST as compared with facility based HIV testing. They reported that since they do the test themselves, they remain with no doubts about the test results. On the other hand, they felt if someone else gave them the results, there was a possibility that results could be altered (intentionally or not). A participant cited an example of false HIV test results they were given at a facility and felt HIVST would be a solution to such a problem.

*“The other thing is that here I will be able to see the results by myself and know that, I am like this. This is how my status is, when I can personally see the results”(M03, Discordant)*.

It therefore appears that HIVST would also possibly eliminate errors of false HIV test results resulting from transcription (recording), as participants are able to check for themselves the results of the test results. This also eliminates mistrust that health workers could provide wrong results intentionally.

Despite its acceptability, participants noted a few concerns regarding HIVST, the common one being the psychological/emotional concerns associated with an HIV positive test result. Participants felt that whereas privacy was an advantage in the HIVST approach, it also stood the challenge of lack of emotional support in case one turned HIV positive. This could also be associated with a lack of post-test counselling in general, even when one turns to be HIV negative, which requires them to be adequately guided on the next steps to maintain their HIV negative status. Others felt that this approach also may result into false results if one panics and ends up misinterpreting the test results.

*“Counselling for this approach is lacking. For example, if you have always known yourself to be HIV negative and you find yourself HIV positive, handling the situation may be difficult when you are alone, with no body to council you”*. (M07, FSW).

However, the emotional concerns were expressed by fewer (six) participants compared with those (14) who felt that was not a problem to them.

### 3.4 Preferred HIV testing method, willingness to pay for the kit, amount one was willing to pay and preferred kits distribution model

**Table 2:** Distribution of respondents by preferred HIV testing method, willingness to pay for the kit, amount one was willing to pay and preferred kits distribution mode.

|  | n | % |
| --- | --- | --- |
| **Preferred HIV testing method (N=361)** |  |  |
| Conventional health facility testing | 31 | 8.6 |
| Blood-based HIVST | 11 | 3.0 |
| Oral fluid-based HIVST | 319 | 88.4 |
| **Willingness to pay for HIVST kit (N=361)** |  |  |
| Yes | 264 | 73.1 |
| No | 97 | 26.9 |
| **Amount to pay (N=361)** |  |  |
| Not willing | 97 | 26.9 |
| Up to 5000 UGX | 226 | 62.6 |
| More than 5000 UGX | 38 | 10.5 |
| **Kits distribution model (N=330)** |  |  |
| Pick from health facility | 108 | 32.7 |
| Home delivered by health workers | 121 | 36.7 |
| Home delivered by peers | 74 | 22.4 |
| Buy from private facility | 27 | 8.2 |

### 3.5 Preferred method for HIV testing during routine follow up monitoring while on PrEP

Majority of respondents (88.4%, n=319) preferred to use an oral fluid based HIVST for routine three monthly HIV testing while on PrEP compared to the conventional health care provider-dependent and facility-based HIV testing (8.6%, n=31). Only 3% (n=11) preferred a blood based self-test kit if introduced in the future.

Participants highlighted various advantages of the oral based HIVST over the other two HIV testing options as being: non-invasive, not painful, no blood loss, and easy to use.

*“I would prefer the oral based test, because for it will not be difficult like the others. The one of a finger prick may be difficult for me to use, because I do not want to be pricked. They prick you and you get damages on your fingers” (M05, MSM/MSW)*.

*“From among the three, this one of oral fluid test is the best, because the other two are painful” (M08, PWUD)*.

Other participants expressed fear of continued blood loss during routine three monthly HIV testing procedures and felt that the continued blood loss could cause them health complications, yet with the oral fluid based test, no such fears could arise.

> *I have found this one (oral fluid based HVST approach) different from others because for me, I don’t want to lose my blood, because sometimes when there are checking you, they take off quite a lot of blood, but here there is no blood I lose and yet I will be sure of the results, just like the ones I will get using the blood test (M02, Discordant)*.

In terms of ease of use, participants found the oral based HIVST easier compared with the other approaches. For example, participants explained that the test procedure was easy for lay persons compared e.g. with the finger prick based HIVST, where the technique of drawing blood could be more difficult. Whereas this concern could result from lack of training in the finger prick approach, some felt even with training, the finger prick would still be more complex as it required one piercing themselves (which many also be feared) as opposed to only paring a test instrument around the mouth, a procedure many compared to brushing of the teeth. Hence, the majority felt the oral fluid based HIVST approach was more suitable to lay people compared with the finger-prick approach.

*“And sometimes you may use it (the finger prick based test) wrongly since you are not a health professional. So, it may not treat you well. But this method is so far the best” (M01, PUWD)*.

### 3.6 Willingness to pay and amount to pay for HIV self-testing kits

Majority of respondents (73.2% (n=264) were willing to pay for an HIV self-testing kit but despite their willingness to pay, they felt kits should be provided free of charge or sold at a very reduced cost and 85.6% (n=226) were willing to pay an amount not exceeding 5000/ Uganda shillings ($1.4).

*“If this kit comes at a cost, it will be difficult for me to pay. Why…because I don’t have a permanent job which I can use to pay. But if it comes free of charge, everyone will want to use it”. (M06, MSM)*.

*“If the test came at a cost, that is a dead deal…it’s a dead deal because not everyone has money. There are those who have, meaning it will be the rich to use them” (M08, PWUD)*.

*“As for me I want it freely. At least if it came at around 2000 ($0*.*6), there I can afford, but still you see that even people fail to afford a pregnancy test, so when you are doing something, you also put into consideration that people fall under different categories” (M10, FSW)*.

### 3.7 Preferred HIV Self-Testing kit distribution approaches/models among Key Populations taking PrEP in central Uganda

Participants expressed views of where they would prefer to receive the oral based HIVST kits as both public and private health facilities, small and large health facilities, community centres and groups, and commercial sites such as shops. Although many distribution models were suggested, the motivation behind the choice was related to the ease of access, cost implications and privacy concerns. For example, some preferred their current health facilities (facility based model), even if these were far from their residencies because they trust their primary service providers. Other participants preferred to receive the kits from their peers (Community Based Model), citing that they find it easy to identify with fellow peers than strangers, while others preferred the kits to be distributed at all levels of health care facilities to facilitate accessibility by all those who need them.

> *We also have our peers who can distribute them to us. Even I am also a peer, so we can get condoms and go and distribute them to the communities. So even this approach can work. They train you and you come to know how to use it. Because for me I know but there is someone who does not know. So, that approach would work, especially for us (MSMs), who don’t want to go to hospital, who can even spend a year, or six months, such people would benefit a lot more (M05, MSM)*.

Participants expressed concerns about cost and integrity of the kits if they were to be distributed at private facilities hence government facilities were more preferred as opposed to private and commercial sites.

*“I don’t want to find it from any other place, because they are many people who duplicate. Like this test, you may find its duplicates by tomorrow. So if the government decides to bring them, we should access them from the main hospital, Mulago, from a qualified doctor” (M08, PWUD)*.

## Discussion

Our study findings reveal that HIVST is highly acceptable among people taking PrEP irrespective of their social demographic and economic characteristics. These findings are comparable to other studies; for instance, acceptability of HIVST was 100% among pregnant women in India [16] and in Kenya, acceptability and feasibility of HIVST among people taking PrEP was found to be 98% [12]. However, unlike our study which involved a variety of KPs and PPs, the study by Sarkar and colleagues [16] was conducted among pregnant women. Similarly, the Kenya study focussed on only KPs on PrEP who were in sero discordant relationships [12]. Our study, having catered for a variety of KPs and PPs including MSM, sero discordant, FSW, PWUIDS, fisher folks, adolescent young girls and women provides grounded evidence on the acceptability of HIV self-testing among respondents taking PrEP. Acceptability results in our quantitative study are further justified by the qualitative findings in which respondents singled out key reasons to justify self-testing as: The procedure being time saving, cost effective (eliminates transport costs), private, and accessible; findings that are similar to those from previous research [17].

Compared to other studies elsewhere, Ng and colleagues [18], in their study to determine accuracy and user acceptability of HIV self-testing using Oraquick HIVST kit, 87.4% of participants were willing to pay for the kit, and out of those willing to pay, only 28% could afford the kit at a market price of five USD. Similarly, Maheswaran and others [19], in their study entitled “Cost and quality of life analysis of HIV self-testing and facility-based HIV testing and counselling in Blantyre” recommended that HIVST would be affordable if the kits were subsidized to a cost comparable to that of routine HIV facility based testing. Our findings further suggest that HIVST test kits should be free of charge or at a very minimal cost that KPs and PPs can afford.

This study indicated that there is no single most popular HIVST kits distribution model that would apply to all categories of key and priority populations. However, three most preferred distribution models were home delivered by health workers, picking from health facility, and home delivered by peers. These findings relate with those from other studies which favored home based [20], facility based, and peer facilitated [5] models. Special groups such as FSWs and fisher folks preferred the peer distribution approach compared to the rest of the KPs, while some groups such as fisher folks also considered accessing the kits at commercial sites such as drug shops for easy access.

Our study had several limitations. A non-probability consecutive convenience sampling method was used to enrol respondents (due to time and logistical constraints plus the dynamic nature of the study population|) hence not all had an equal chance of being selected. This could have led to a selection bias.

By providing test kits to respondents and requesting them to test on the same day could have influenced their decision to accept self-testing without adequately reflecting on their choice. A better method if time could allow, would have been to let respondents go with the kits, and decide to or not to perform self-testing. The Oral fluid HIV testing kits used in this study could miss early HIV infections that can be detected by other HIV blood-based platforms especially in populations with high HIV incidence such as KPs and PPs where acute infection is more common hence it is possible that some early infections could not be detected.

## Conclusions

HIV self-testing is highly acceptable and is the most preferred HIV testing approach for routine three monthly follow up testing among KPs and PPs on PrEP in central Uganda. The most preferred models of HIVST kt distribution are self-pick from facility, heath worker distribution and peer distribution. Oral fluid-based HIV self-testing kits should be provided free or subsidised to not more than 5,000 Uganda shillings (USD 1.4) since majority of KPs and PPs will not afford the kits beyond this price. We recommend that: HIVST be adopted as a testing approach for all people who may require regular rapid HIV testing due to ongoing risk of exposure on opt-out basis; Efforts should be made to provide adequate pre and post “self-testing” counselling information to all people who intend to perform self-testing in order to eliminate possible psychological stress arising from self-testing results. There is need to study linkage to care after HIV self-testing as this was not addressed by our study but was a concern during our qualitative assessment.

## Data Availability

Deidentified data will be made available by contacting the study PI, but is also available at https://doi.org/10.7910/DVN/XLBZXD

## Acknowledgements

We thank the study participants for their time. We also acknowledge the staff at the two study sites, including nurses, counsellors and clinicians for their work on this study. We thank Ministry of Health Uganda for providing guidance and linking us to the study sites.

## References

1. UNAIDS. Global HIV & AIDS statistics — Fact sheet 2022 [Available from: https://www.unaids.org/en/resources/fact-sheet.

2. WHO. Guideline on when to start antiretroviral therapy and on pre-exposure prophylaxis for HIV. Geneva: World Health Organization 2017.

3. MoH. Technical Guidance on Pre-Exposure Prophylaxis (PrEP) for Persons at High Risk of HIV in Uganda. In: Health, editor. Uganda: Ministry of Health; 2016.

4. UNAIDS. A short technical update on self-testing for HIV. United Nations Program on HIV/AIDS; 2014.

5. Geng EH, Ortblad K, Kibuuka Musoke D, Ngabirano T, Nakitende A, Magoola J, et al. Direct provision versus facility collection of HIV self-tests among female sex workers in Uganda: A cluster-randomized controlled health systems trial. PLoS medicine. 2017;14(11):e1002458.

6. Nangendo J, Obuku EA, Kawooya I, Mukisa J, Nalutaaya A, Musewa A, et al. Diagnostic accuracy and acceptability of rapid HIV oral testing among adults attending an urban public health facility in Kampala, Uganda. PLoS One. 2017;12(8):e0182050.

7. Mugo PM, Micheni M, Shangala J, Hussein MH, Graham SM, Rinke de Wit TF, et al. Uptake and Acceptability of Oral HIV Self-Testing among Community Pharmacy Clients in Kenya: A Feasibility Study. PLoS One. 2017;12(1):e0170868.

8. Kurth AE, Cleland CM, Chhun N, Sidle JE, Were E, Naanyu V, et al. Accuracy and Acceptability of Oral Fluid HIV Self-Testing in a General Adult Population in Kenya. AIDS Behav. 2016;20(4):870–9.

9. Katz DA, Golden M R.,, Hughes JP, Farquhar C, Stekler JD, editors. Acceptability and Ease of Use of Home Self-Testing for HIV among Men Who Have Sex with Men 2012; University of Washington.

10. Pal K, Ngin C, Tuot S, Chhoun P, Ly C, Chhim S, et al. Acceptability study on HIV self-testing among transgender women, men who have sex with men, and female entertainment workers in Cambodia: A qualitative analysis. PLOS One. 2016;11(11).

11. Figueroa C, Johnson C, Verster A, Baggaley R. Attitudes and Acceptability on HIV Self-testing Among Key Populations: A Literature Review. AIDS and Behavior. 2015;19(11):1949–65.

12. Ngure K, Heffron R, Mugo N, Thomson KA, Irungu E, Njuguna N, et al. Feasibility and acceptability of HIV self-testing among pre-exposure prophylaxis users in Kenya. J Int AIDS Soc. 2017;20(1):21234.

13. Walensky RP, Bassett IV. HIV self-testing and the missing linkage. PLoS Med. 2011;8(10):e1001101.

14. UNCST. National Guidelines for Research involving Humans as research Participants. National Guidelines for Research involving Humans as research Participants. Kampala: UNCST website; 2014. p. 38.

15. John Santelli SH, Terry McGovern. Inclusion with Protection: Obtaining informed consent. UNICEF Innocenti Research Broef. 2017:13.

16. Sarkar A, Mburu G, Shivkumar PV, Sharma P, Campbell F, Behera J, et al. Feasibility of supervised self-testing using an oral fluid-based HIV rapid testing method: a cross-sectional, mixed method study among pregnant women in rural India. Journal of the International AIDS Society. 2016;19(1):20993.

17. Weiser SD, Pant Pai N, Sharma J, Shivkumar S, Pillay S, Vadnais C, et al. Supervised and Unsupervised Self-Testing for HIV in High- and Low-Risk Populations: A Systematic Review. PLoS Medicine. 2013;10(4):e1001414.

18. Ng OT, Chow AL, Lee VJ, Chen MI, Win MK, Tan HH, et al. Accuracy and user-acceptability of HIV self-testing using an oral fluid-based HIV rapid test. PLoS One. 2012;7(9):e45168.

19. Maheswaran H, Petrou S, MacPherson P, Choko AT, Kumwenda F, Lalloo DG, et al. Cost and quality of life analysis of HIV self-testing and facility-based HIV testing and counselling in Blantyre, Malawi. BMC Medicine. 2016;14(1):34.

20. Gaydos CA, Hsieh YH, Harvey L, Burah A, Won H, Jett-Goheen M, et al. Will patients “opt in” to perform their own rapid HIV test in the emergency department? Ann Emerg Med. 2011;58(1 Suppl 1):S74–8.

